# Risk of cardiovascular events following COVID-19 in people with and without pre-existing chronic respiratory disease

**DOI:** 10.1101/2023.03.01.23286624

**Authors:** Hannah Whittaker, Constantinos Kallis, Thomas Bolton, Angela Wood, Samantha Walker, Aziz Sheikh, Alex Brownrigg, Ashley Akbari, Kamil Sterniczuk, Jennifer Quint, CVD-COVID-UK/COVID-IMPACT Consortium

## Abstract

**Background:** COVID-19 is associated with a higher risk of cardiovascular outcomes in the general population, but it is unknown whether people with pre-existing chronic respiratory disease (CRD) have a higher risk of cardiovascular events post-COVID-19 compared with the general population and, if so, what respiratory-related risk factors may modify this risk in these people.

**Methods:** Primary and secondary care data from the National Health Service and COVID-19-specific linked data were used to define a population of adults in England with COVID-19 (index date) between 01/01/2020-30/11/2021. Adjusted Cox Proportional Hazard regression was used to quantify the association between CRD, asthma-related factors, COPD-related factors, and risk of cardiovascular events. CRD included asthma, COPD, bronchiectasis, cystic fibrosis, or pulmonary fibrosis prior to COVID-19 diagnosis. Asthma-specific factors included baseline asthma control, exacerbations, and inhaled corticosteroid (ICS) dose. COPD-specific risk factors included baseline ICS prescriptions and exacerbations. Secondary objectives quantified the impact of COVID-19 hospitalisation and vaccine dose on cardiovascular outcomes.

**Results:** Of 3,670,455 people, those with CRD had a modest higher risk of cardiovascular events (HR_adj_ 1.11, 95%CI 1.07-1.14), heart failure (HR_adj_ 1.15, 1.09-1.21), and pulmonary emboli (HR_adj_ 1.20, 1.11-1.30) compared with people without CRD. In people with asthma, baseline exacerbations and high-dose ICS were associated with a higher risk of cardiovascular outcomes (HR_adj_ 1.24, 1.15-1.34 and 1.12, 1.01-1.24, respectively). In people with COPD, exacerbations were associated with a higher risk of cardiovascular outcomes (HR_adj_ 1.40, 1.28-1.52). Regardless of CRD, the risk of cardiovascular events was lower with increasing COVID-19 vaccine dose.

**Conclusions:** Higher risk of cardiovascular events following COVID-19 might be explained at least in part by the underlying CRD and severity of that condition. In addition, COVID-19 vaccines were beneficial to both people with and without CRD with regards to CV events.

**Key Messages:** Pre-existing chronic respiratory disease, asthma and COPD severity were associated with a higher risk of various types of cardiovascular outcomes following COVID-19. Regardless of having pre-existing chronic respiratory disease, COVID-19 vaccination reduced the risk of cardiovascular events following COVID-19.

## Introduction

The SARS-CoV-2 virus induces a prothrombotic and proinflammatory state, which increases the risk of thrombotic and inflammatory conditions (1). These processes can lead to the development of chronic and acute diseases, including chronic and acute cardiovascular events such as coronary artery disease, myocardial infarction (MI), stroke, and venous thromboembolism (VTE) (2, 3, 4, 5, 6). One group of people in whom the risk of cardiovascular disease is higher than in the general population is those with chronic respiratory disease (7, 8). Previous studies have shown that in people with chronic obstructive pulmonary disease (COPD), risk of cardiovascular events increases, in particular following an acute pulmonary infection such as an exacerbation of COPD (9). Therefore, it is possible that people with chronic respiratory disease may have a different risk of developing cardiovascular outcomes following an infection such as COVID-19, compared with people without chronic respiratory disease.

Cardiovascular disease is a common comorbidity in people with COPD, asthma, interstitial pulmonary fibrosis, and bronchiectasis (7, 10) (11, 12, 13) (14, 15) (16). Whilst there is less extensive literature on cystic fibrosis, evidence suggests that people with cystic fibrosis have a high prevalence of risk factors for cardiovascular disease (17). Given that chronic respiratory diseases are the third leading cause of death worldwide and are associated with significant morbidity, it is important to better understand the long-term impact of COVID-19 to improve the management of people with chronic respiratory disease post-COVID-19 (10). In addition, it remains unknown as to whether the association between COVID-19 vaccination and the reduced risk of cardiovascular events remains in people with and without chronic respiratory disease separately, and to what extent this risk differs between those with and without chronic respiratory disease given that people with chronic respiratory disease already have a different baseline level of inflammation(6, 18) (7). Studies have also demonstrated that people with more severe chronic respiratory disease are more likely to develop cardiovascular events therefore, it is important that we better understand whether people with severe respiratory disease have an increased risk of cardiovascular events following COVID-19 compared to those with mild or moderate disease severity (19).

Using routinely collected electronic health record (EHR) data sources in England, we investigated the association between pre-existing chronic respiratory disease and cardiovascular outcomes after COVID-19 in a population of people with COVID-19 in the community and for people hospitalised for COVID-19. We then investigated the association between markers of asthma and COPD disease severity and risk of cardiovascular events following COVID-19 as asthma and COPD are the leading causes of chronic respiratory disease worldwide (10). Finally, we aimed to investigate the association between COPD exacerbations, ICS use and cardiovascular events following COVID-19.

## Methods

### Study population

Pseudonymized data used in this study were accessed and analysed through the British Heart Foundation Data Science Centre CVD-COVID-UK/COVID-IMPACT consortium within the NHS Digital secure, privacy-protecting TRE Service for England (20). The data included: routinely collected primary care data, hospital data (including inpatient, outpatient, emergency department, and critical care), mortality data (including causes of death from death certificates), COVID-19 laboratory test data, COVID-19 vaccine data, and data on dispensed medications (Table E1). All code lists for variables used int his study can be found: https://github.com/BHFDSC/CCU035_01/blob/main/phenotypes/CCU035_01-D01-codelist.py.

Primary care data from General Practice Extraction Service (GPES) for Pandemic Planning and Research (GDPPR) included over 56 million people registered at a general practice in England who were alive on 30^th^ November 2019. Age, sex, general practice, and COVID-19 diagnosis variables were used to define a population of people aged 18 years or older who had been diagnosed with COVID-19 since 1^st^ January 2020. COVID-19 diagnosis was determined through primary care records, hospital records and SARS-CoV-2 laboratory test data. People were excluded if they had a COVID-19 vaccination before their first COVID-19 diagnosis. Index date was defined as the first COVID-19 diagnosis date on or after 1^st^ January 2020. People were followed up until 30^th^ November 2021, or earlier at individual’s second COVID-19 diagnosis date, first COVID-19 vaccination date, or the date at which individuals died (Figure E1).

Data quality checks were implemented to ensure data validity. Specifically, we ensured that: 1) date of birth was recorded before date of death; 2) recorded sex and date of birth were not missing; and 3) there were no recorded pregnancies or births for men and no recorded prostate cancer for women.

### Pre-existing respiratory disease

Pre-existing chronic respiratory disease was defined as having a primary care diagnosis of asthma, COPD, bronchiectasis, cystic fibrosis, or pulmonary fibrosis at any point prior to index date (i.e., first COVID-19 diagnosis date). The groups of pre-existing respiratory diseases were not mutually exclusive.

### Asthma cohort

From this base population, a cohort of people with pre-existing asthma was defined. This cohort was used to investigate the association between asthma-specific exposures and post-COVID-19 cardiovascular outcomes. Asthma-related exposures included: asthma exacerbations, asthma control, and asthma severity as defined by ICS dose.

Specifically, asthma exacerbations were defined as at least one of the following criteria: i) a course of oral corticosteroids; ii) a hospital admission for an asthma exacerbation (using the international classification of diseases (ICD)-10 codes in any position); and iii) an emergency department visit for an asthma-related cause.

Asthma control was defined as at least one of the following criteria: i) two or more courses of oral corticosteroids or two or more hospital admission or accident and emergency attendances in the year prior to index; ii) at least six months of short-acting beta agonist (SABA) use in the year prior to index date; iii) forced expiratory volume in 1 second (FEV1)/forced vital capacity (FVC) less than 80% predicted.

Asthma severity was defined using ICS dose based on the global initiative for asthma (GINA) guidelines (21). ICS dose was categorised as low, medium, and high using the last ICS prescription in the year prior to index date.

### COPD cohort

From the base population, a cohort of people with pre-existing COPD was defined. This cohort was used to investigate the association between COPD-specific exposures and post-COVID-19 cardiovascular outcomes. The exposures of interest in this cohort were ICS use and COPD exacerbations.

ICS use was defined in the year prior to index date. Specifically, ICS was defined as having at least one prescription of ICS in 3 of the 4 quarters of the year prior to index date, following previous studies (22). Patients were categorised as ICS users or non-ICS users as the exposure.

COPD exacerbations were defined in the year prior to index date. COPD exacerbations were defined following a previously validated definition in electronic health care records where data were available (23, 24). We defined COPD exacerbations as at least one exacerbation requiring hospitalisation or a prescription of respiratory-related antibiotics and oral corticosteroids for a length of 5-14 days.

### Cardiovascular outcomes

Cardiovascular (CV) outcomes were defined using primary care records, secondary care data, and mortality data. Cardiovascular events were determined from the start of follow-up to the end of follow-up. The first cardiovascular event during follow-up was used. Cardiovascular outcomes were defined as the following groups: i) VTE events, including deep vein thrombosis and portal vein thrombosis; ii) coagulopathy events, including disseminated intravascular coagulopathy, thrombotic thrombocytopenic purpura, thrombocytopenia and thrombophilia; iii) heart failure (HF), including HF and cardiomyopathy; iv) angina, including both stable and unstable angina; v) myocarditis and pericarditis; vi) stroke, including ischaemic and haemorrhagic strokes; vii) MI; viii) pulmonary embolism; and ix) arrhythmias. A composite cardiovascular outcome was also defined as a combination of all groups of cardiovascular events. For the asthma and COPD cohorts, these outcomes events were categorised into four variables: composite CV, arterial CV, venous CV, and other CV events due to smaller population numbers. Arterial CV events included disseminated intravascular coagulation-related events, stroke-related events, and myocardial infarction. Venous CV events included venous thrombosis-related events and pulmonary embolism-related events. Other CV events included HF, angina, myocarditis, pericarditis, and arrhythmias.

### COVID-19 vaccination

COVID-19 vaccination date and dose number data were extracted from the National Immunisation Management System COVID-19 vaccination database. Where dose number did not match up with vaccine date, dose number was recalculated to align with dates at which people were vaccinated. In addition, all dates were checked to ensure no duplication and vaccine dates that were within follow-up were included.

### Statistical analyses

Our analysis was performed according to a pre-specific analysis plan published on GitHub, along with the phenotyping and analysis code (https://github.com/BHFDSC/CCU035_01). Cox Proportional Hazard regression was used to quantify the association between pre-existing respiratory disease and risk of cardiovascular outcomes following COVID-19. Standard error estimates were adjusted for clustering of people in general practice as it was possible the treatment of people with COVID-19 differed by general practices across England. Cox regression was also used to investigate the association between i) asthma or COPD and post-COVID-19 cardiovascular events in the base population; ii) asthma exacerbations, asthma control, and ICS dose and post-COVID-19 cardiovascular events in the asthma cohort; and iii) ICS use and post-COVID-19 cardiovascular events in the COPD cohort. Fully adjusted models were adjusted for the following baseline covariates: age, gender, smoking status, obesity, surgery in the last year, a history of hypertension, diabetes, cancer, liver disease, dementia, chronic kidney disease, depression, CVD, CVD-related medications 3 months prior to index date, region, index of multiple deprivation, and ethnicity. Additional COPD and asthma-specific covariates included exacerbations, gastro-oesophageal reflux disease, ICS, dyspnoea, and COPD duration. Missing data were reported, and complete case analysis was used.

Secondary analyses for the main analysis included the following:

i) Effect modification by COVID-19 severity determined by whether people were hospitalised for COVID-19 within 2 weeks of their COVID-19 diagnosis.
ii) Effect modification by vaccination status whereby follow-up time was split into four possible time periods: the unvaccinated period (reference category), the period from 1^st^ vaccine to 2^nd^ vaccine, the period from 2^nd^ vaccine to 3^rd^ vaccine (booster vaccination), and the period from the 3^rd^ vaccine to the end of follow-up. All types of COVID-19 vaccins were used. In this analysis end of follow-up was defined as 30/11/2021, second COVID-19 diagnosis, or death. Cardiovascular events from the index date to the end of follow-up were identified.

A sensitivity analysis was performed whereby people with a history of acute cardiovascular outcomes (venous thrombosis, dissected artery, and stroke) event prior to COVID-19 diagnosis were excluded. Due to multiple testing, Bonferroni correction was applied whereby for each set of models in the main analysis, a p-value threshold of 0.005 was used. For the asthma and COPD-specific analyses, a p-value threshold of 0.0125 was used (see supplement p1). To ensure anonymity and compliance with the CVD-COVID-UK consortium, all reported numbers were rounded to the nearest 5. Counts less than 10 were expressed as “<10”.

## Results

A total of 3,670,460 people were diagnosed with COVID-19 between the 1^st of^ January 2020 and the 30^th^ November 2021 and before their first COVID-19 vaccination date. Of these, 670,055 (18.3%) had pre-existing chronic respiratory disease and 3,000,405 (81.7%) had no pre-existing chronic respiratory disease (Figure 1). A total of 606,980 (16.5%) had asthma, 83,515 (2.3%) had COPD, 17,095 (0.5%) had bronchiectasis, and 8,980 (0.2%) had pulmonary fibrosis, and 715 (0.02%) had cystic fibrosis. The groups of pre-existing chronic respiratory diseases were not mutually exclusive. Median follow-up from COVID-19 diagnosis to end of follow-up, including censoring at the date of a composite cardiovascular event, death, COVID-19 vaccination, and 2^nd^ COVID-19 diagnosis, was 3.9 months (IQR 1.9 to 6.6 months). A total of 46,255 (1.3%) people had a recorded cardiovascular event during the follow-up. Table E2 reports the number of events that occurred for each cardiovascular outcome.

**Figure 1:**
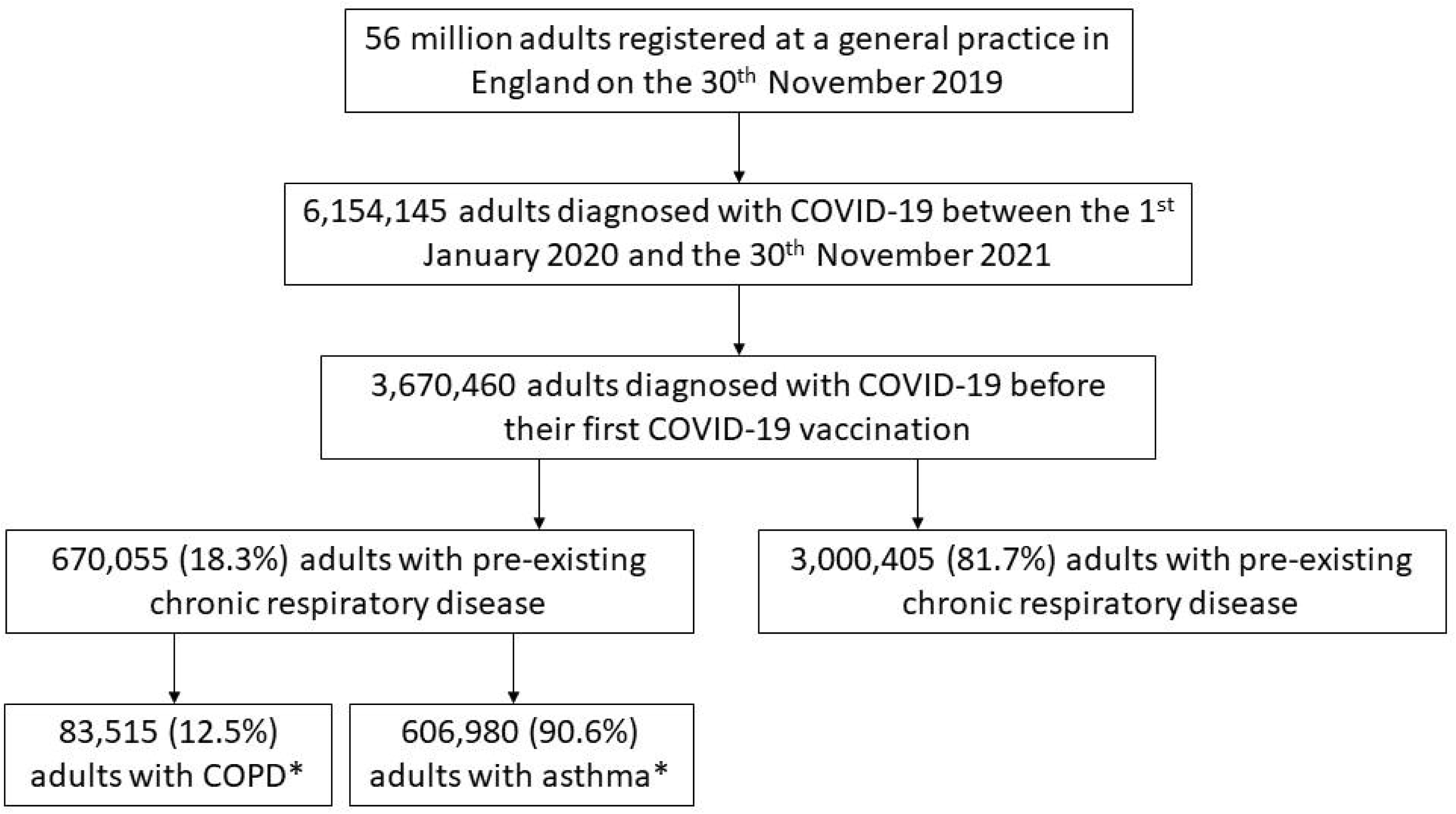
Flow diagram illustrating inclusion of study population.

People with and without pre-existing respiratory disease were similar in terms of all baseline characteristics however, those with pre-existing respiratory disease were more likely to be ex-smokers, less likely to live in London, were more likely to be obese, had surgery in the last year, have hypertension or chronic kidney disease, and have cardiovascular disease prior to COVID-19 diagnosis (Table E3).

### Risk of cardiovascular events in people with pre-existing respiratory disease compared with people without pre-existing respiratory disease post-COVID-19 diagnosis

People with pre-existing respiratory disease had an overall higher risk of cardiovascular outcomes compared with those without pre-existing respiratory disease (adjusted HR 1.06, 95% CI, 1.03-1.08). Specifically, people with pre-existing respiratory disease had a higher risk of HF (adjusted HR 1.17,

95% CI 1.13-1.22) and pulmonary embolism (adjusted HR 1.20, 95% CI 1.12-1.29) compared with those without pre-existing respiratory disease and were less likely to have a stroke event compared to those with pre-existing respiratory disease (adjusted HR 0.88, 95% CI 0.84-0.92) (Figure 2, Table E4).

**Figure 2:**
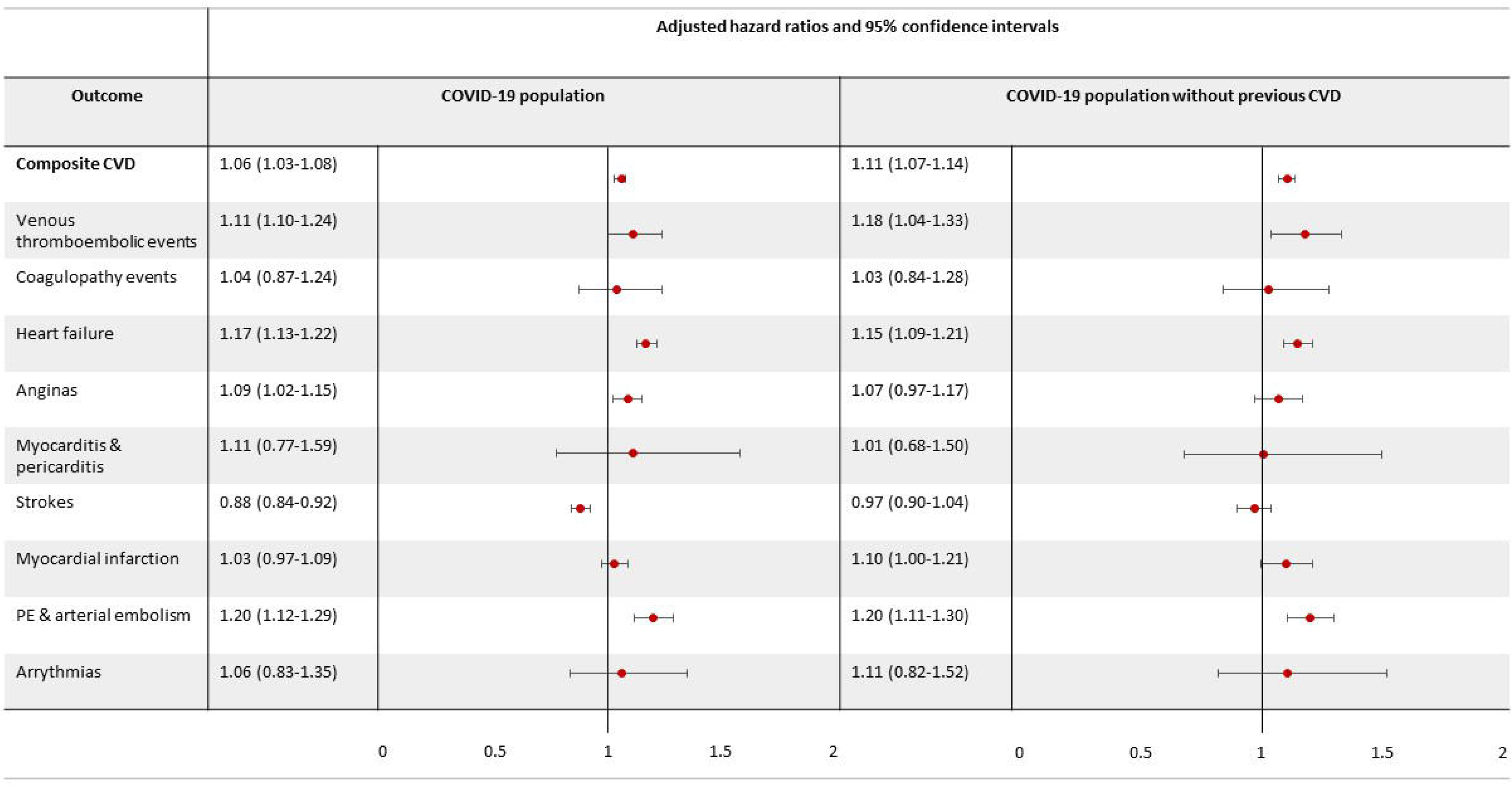
Risk of post-COVID-19 cardiovascular outcomes (and each cardiovascular component) in people with pre-existing chronic respiratory disease compared with people without pre-existing chronic respiratory disease. Legend: Venous thromboembolic events include deep vein and portal vein thrombosis. Coagulopathy events include thrombocytopenia, thrombophilia and mesenteric thrombus. PE (pulmonary embolism). Risk of cardiovascular outcomes is reported in all people with COVID-19 and people with no history of cardiovascular events prior to COVID-19 separately.

After excluding people with a history of severe cardiovascular prior to COVID-19, people with pre-existing respiratory disease had a higher risk of any cardiovascular event (adjusted HR 1.11, 95% CI 1.07-1.14), HF (adjusted HR 1.15, 95% CI 1.09-1.21) and pulmonary embolism (adjusted HR 1.20, 95% CI 1.11-1.30) compared with people with no pre-existing respiratory disease. People with and without pre-existing respiratory disease had a similar risk of stroke. Supplementary Table E5 report crude and fully adjusted estimates.

### Effect of COVID-19 severity on risk of cardiovascular outcomes in people with and without pre-existing respiratory disease

Out of 3,670,460 people who had COVID-19, 250,370 (6.8%) were hospitalised compared with 3,420,085 (93.2) who were not. In people who were not hospitalised for COVID-19, there was an association between pre-existing chronic respiratory disease a higher risk of composite cardiovascular disease (adjusted HR 1.15 95% CI 1.11-1.19) including HF and pulmonary embolisms similar to the main results. However, in people who were hospitalised for COVID-19, there was an association between pre-existing chronic respiratory disease and decreased risk of stroke only (adjusted HR 0.79, 95% CI 0.69-0.90) (Tables E6-E7).

### Effect of COVID-19 vaccination on risk of cardiovascular outcomes in people with and without pre-existing respiratory disease

A total of 2,652,210 (70.4%) people had at least one COVID-19 vaccine after their first COVID-19 diagnosis up until 30^th^ November 2021. Of these people, 218,970 (8.3%) had 1 vaccine, 1,708,275 (64.4%) had 2 vaccines, and 724,965 (27.3%) had the two COVID-19 vaccines and the COVID-19 booster. Compared with the period between COVID-19 diagnosis and dose 1 (i.e., the unvaccinated period), the risk of composite cardiovascular outcomes was lower with each subsequent COVID-19 vaccine dose. There was no effect modification of pre-existing respiratory disease on the association between dose number and risk of cardiovascular outcomes (Figure 3, Table E8).

**Figure 3:**
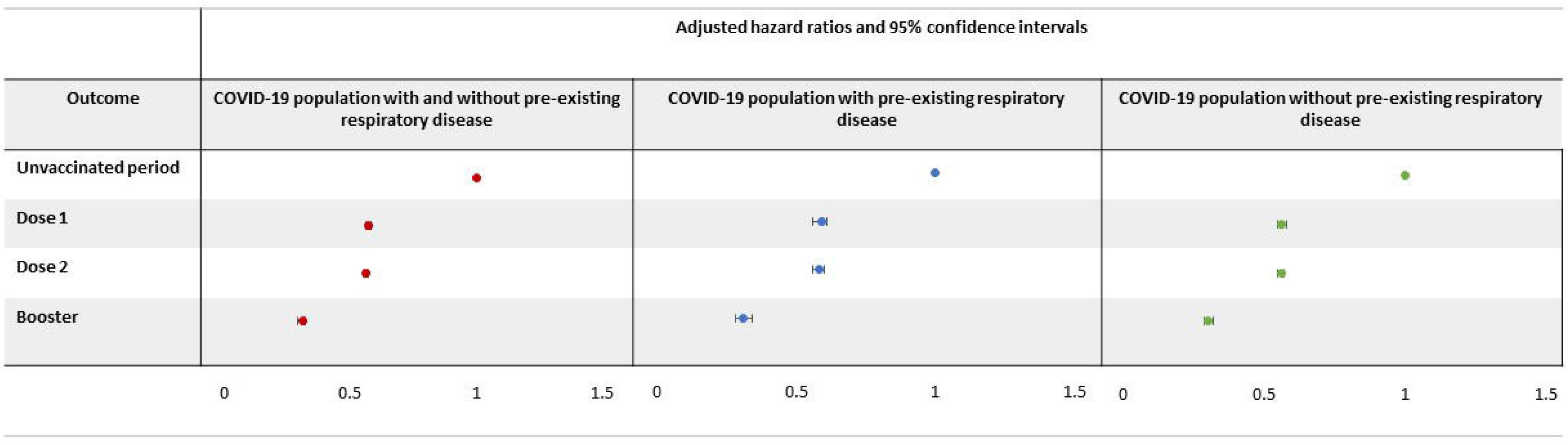
Association between increasing number of COVID-19 vaccinations and risk of cardiovascular outcomes in people who had been diagnosed with COVID-19. Legend: Reference period is the unvaccinated period from COVID-19 diagnosis to first COVID-19 vaccination or censoring. The dose 1 period is from 1^st^ COVID-19 vaccination to 2^nd^ COVID-19 vaccination or censoring. The dose 2 period is from the 2^nd^ COVID-19 vaccination to booster or censoring. The booster period is from the booster vaccine to end of follow-up or censoring.

### Asthma and risk of CV outcomes following COVID-19

People with pre-existing asthma also had an overall higher risk of composite cardiovascular events as well as heart failure and pulmonary emboli compared to people without pre-existing asthma (adjusted HR 1.04, 95% CI 1.01-1.07, adjusted HR 1.12, 95% CI 1.07-1.17, adjusted HR 1.16, 95% CI 1.07-1.26, respectively; Table E9) and were less likely to have a stroke event compared with those with pre-existing asthma (adjusted HR 0.87, 95% CI 0.83-0.92). After excluding patients with a history of CVD prior to COVID-19, a similar pattern of association was seen however, there was no association between pre-existing asthma and stroke.

In addition, of the 606,980 people with asthma, patients with better asthma control had a modest higher risk of composite CV outcomes and arterial events compared with people with poorer asthma control (adjusted HR 1.10, 95% CI 1.03-1.18 and 1.18, 1.05-1.33, respectively; figure 4 and table E10). However, this association disappeared after excluding people with a history of CV disease. In addition, people with asthma with at least one exacerbation at baseline had a higher risk of composite CV outcomes, venous-related outcomes, and other CV-related outcomes compared with people with asthma who did not exacerbate at baseline (adjusted HR 1.24, 95% CI 1.15-1.34, 1.18, 1.05-1.33, 1.28, 1.16-1.42, respectively). After excluding people with a history of CV disease, people with asthma who exacerbated at baseline at a higher risk of composite CV outcomes, venous-related outcomes, and other CV outcomes, but not arterial-related outcomes. Lastly, people with asthma on high dose ICS had a higher risk of other CV related events following COVID-19 compared with people on low dose ICS in people with asthma with and without a history of CV disease (adjusted HR 1.32, 95% CI 1.15-1.52, and 1.29, 1.13-1.49). ICS dose was not associated with arterial or venous CV outcomes.

**Figure 4:**
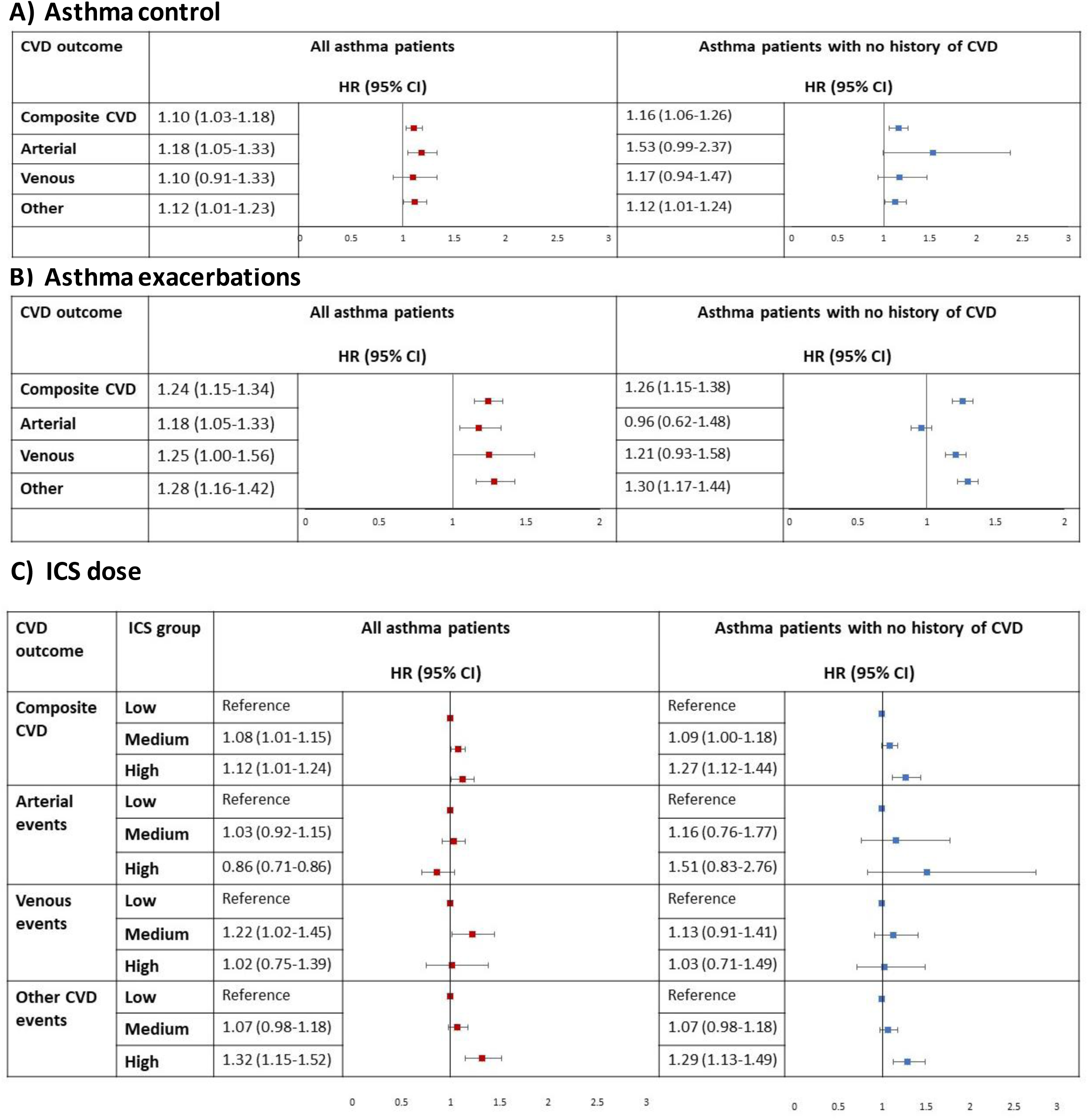
Risk of CV outcomes following COVID-19 by asthma control, asthma exacerbations, and ICS dose in people with asthma. Legend: Arterial events include disseminated intravascular coagulation-related events, stroke-related events, and myocardial infarction. Venous events include venous thrombosis-related events and pulmonary embolism-related events. Other CV events include heart failure, angina, myocarditis, pericarditis, and arrythmias. Risk of CV is reported in all patients with COVID-19 and asthma and asthma patients with no history of CV events prior to COVID-19 separately.

### COPD and risk of CV outcomes following COVID-19

People with pre-existing COPD had an overall increased risk of CV outcomes as well as heart failure and pulmonary emboli compared to those without pre-existing COPD (adjusted HR 1.09, 95% CI, 1.06-1.13, adjusted HR 1.27, 95% CI 1.20-1.33, adjusted HR 1.34, 1.19-1.51, respectively; Table E11) and were less likely to have a stroke event compared with those with pre-existing COPD disease (adjusted HR 0.86, 0.80-0.92; p<0.001). After excluding patients with a history of CVD prior to COVID-19, a similar pattern of association was seen however, there was no association between pre-existing COPD and risk of stroke.

In the population of 83,515 people with COPD, no association between baseline ICS use and risk of venous and other CV related outcomes following COVID-19 was seen however, patients on ICS at baseline had a lower risk of arterial events following COVID-19 compared with patients not on ICS (adjusted HR 0.86, 95% CI 0.76-0.97: figure 5 and table E12). After excluding people with a history of CVD, this association was not seen. On the other hand, COPD patients who exacerbated at baseline had a higher risk of composite CV events, specifically venous and other CV-related events following COVID-19 compared with people who did not exacerbate at baseline in all COPD patients (adjusted HR 1.40, 95% CI 1.28-1.52, 1.67, 1.24-2.24, and 1.53, 1.36-1.72, respectively). These associations remained consistent after excluding people with a history of CVD.

**Figure 5:**
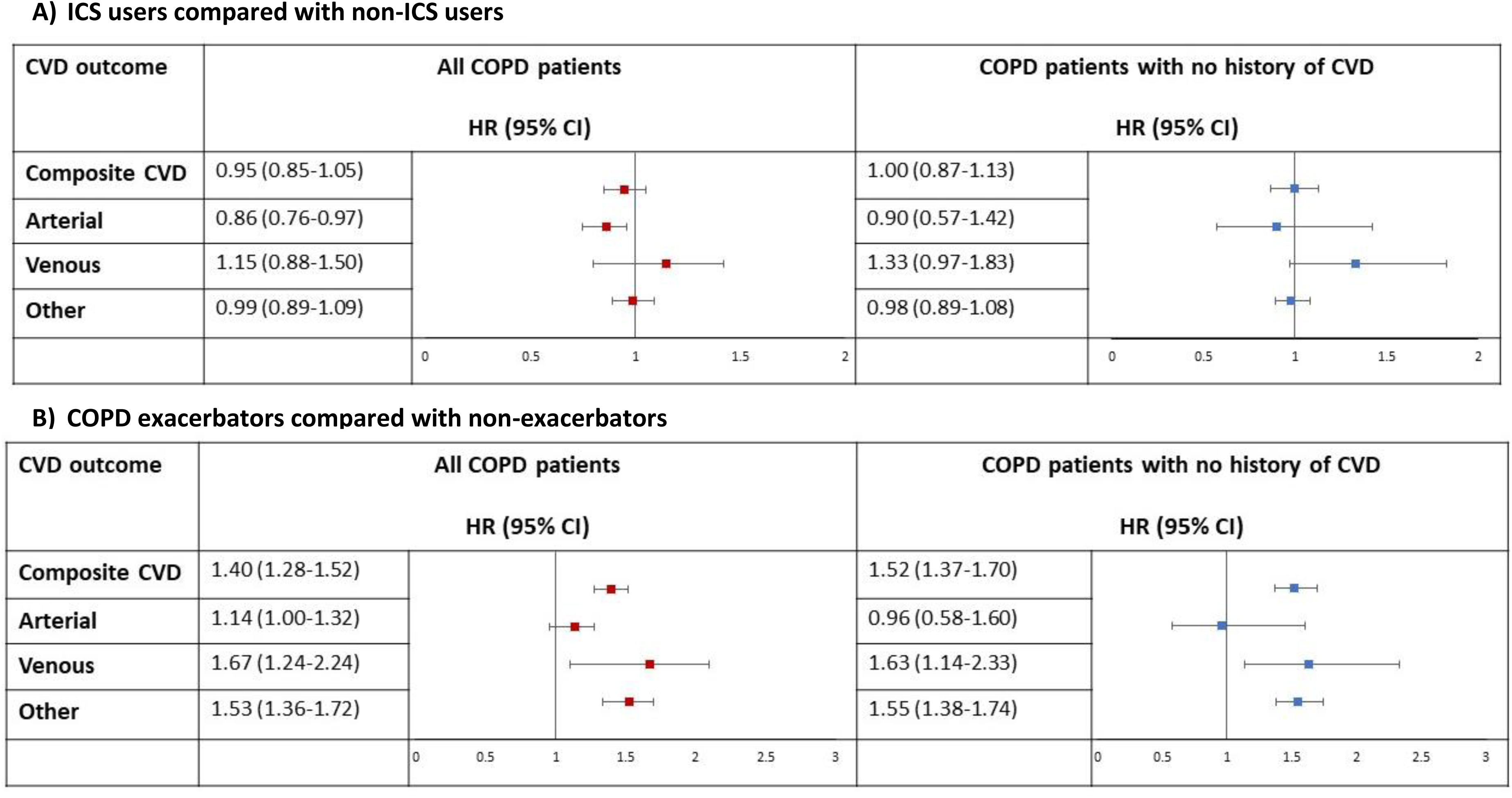
Risk of CV outcomes following COVID-19 by ICS use and COPD in people with COPD. Legend: Arterial events include disseminated intravascular coagulation-related events, stroke-related events, and myocardial infarction. Venous events include venous thrombosis-related events and pulmonary embolism-related events. Other CV events include heart failure, angina, myocarditis, pericarditis, and arrythmias. Risk of CV is reported in all patients with COVID-19 and COPD and COPD patients with no history of CV events prior to COVID-19 separately.

## Discussion

Using routinely collected EHR data on over three million people with COVID-19, we found that having pre-existing chronic respiratory disease was associated with a modest higher risk of cardiovascular events. Specifically, people with pre-existing chronic respiratory disease had a higher risk of heart failure and pulmonary embolism compared with those without pre-existing respiratory disease prior to COVID-19. Additionally, we found that severity of COVID-19, as defined by whether people were hospitalised or not, modified the association between pre-existing respiratory disease and risk of cardiovascular outcomes. In people with COVID-19 who were not hospitalised, those with pre-existing chronic respiratory disease had a higher risk of heart failure and pulmonary embolism compared with those without pre-existing respiratory disease. However, in people with COVID-19 who were hospitalised, those with pre-existing chronic respiratory disease had a reduced risk of stroke and a similar risk of all other cardiovascular outcomes compared with people without pre-existing respiratory disease. In addition, we found that the COVID-19 vaccine reduced the risk of any cardiovascular outcome in people who had been diagnosed with COVID-19 with and without pre-existing respiratory disease to a similar extent. Lastly, we found that in people with asthma or COPD, risk factors associated with an increased risk of cardiovascular events included exacerbations and asthma control and high ICS dose for people with asthma.

Many studies have investigated the risk of cardiovascular outcomes after COVID-19 in the general population. A previous study of 48 million people in England and Wales found that people with COVID-19 had a higher risk of arterial thrombosis and VTE compared with people without COVID-19 and that the risk attenuated with time since COVID-19 diagnosis (5). Other studies have found that people who were hospitalised with COVID-19 had a higher risk of heart failure, stroke, ischemic heart disease, and VTE compared with people who did not have COVID-19 (6, 25, 26). Our study extends previous work by comparing these risks between subgroups of people with pre-existing chronic respiratory disease. A higher risk of heart failure and pulmonary embolism was seen in people with pre-existing respiratory disease. Whilst we adjusted for a history of severe cardiovascular disease, the higher risk of heart failure seen may be due the chronic respiratory disease rather than COVID-19 given HF is a common comorbidity amongst people with chronic respiratory disease with overlapping symptoms (7, 27, 28). Previous studies have shown that the risk of pulmonary embolism is higher in people with asthma and COPD compared with the general population and is particularly higher in people with severe asthma compared with people with mild-to-moderate asthma and during or following an acute exacerbation of COPD (29, 30). Pulmonary embolism and chronic respiratory disease share common risk factors including age and immobilisation however, the exact mechanism driving the association between pulmonary embolisms and chronic respiratory disease is unclear (31). As with heart failure, it is possible that the higher risk of pulmonary embolism in people with pre-existing chronic respiratory disease is due to underlying mechanisms related to respiratory disease or acute infection generally rather than COVID-19.

Surprisingly, we found that people who were hospitalised for COVID-19 (i.e., those with severe COVID-19) with pre-existing respiratory disease had a lower risk of stroke compared with people without pre-existing chronic respiratory disease, but similar risks of all other cardiovascular outcomes. It is possible that the level of COVID-19 severity has a larger impact on cardiovascular outcomes than pre-existing chronic respiratory disease which is why the risk of most cardiovascular outcomes between those with and without chronic respiratory disease was similar in this population. The lower risk of stroke in could be due to better monitoring of people with pre-existing chronic disease in hospital. It is well known that risk of ischemic events increases after acute respiratory events, notably hospitalisation due to acute respiratory events such as exacerbations (9). It is possible that care of people with pre-existing respiratory disease upon hospitalisation with COVID-19 is better because respiratory disease is a known risk factor for stoke.

A previous study found that there was no association between pre-existing cardiovascular disease and risk of cardiovascular outcomes 14 days after hospitalisation in a population of people hospitalised for COVID-19 (32). However, it is possible that this association varies by other pre-existing chronic comorbidities. It is possible that people with pre-existing respiratory disease who had underlying cardiovascular disease prior to COVID-19 were more likely to monitored more efficiently compared with people who did not have pre-existing respiratory disease. For example, having already been prescribed cardiovascular treatment thus reducing risk.

We additionally found that the COVID-19 vaccine lowered the risk of composite cardiovascular outcomes in people with and without pre-existing respiratory disease to a similar extent. Previous studies found that having at least one COVID-19 vaccine post-COVID-19 decreased the rate of various long-covid symptoms including chest pain or tightness, palpitations, and breathlessness (6). Similarly, other studies have found an association between the COVID-19 vaccine and a reduced risk of cardiovascular events, including MI and stroke, after SARS-CoV-2 infection (18).

Furthermore, we found that people with more severe asthma or COPD, as indicated by whether people exacerbated, were on high dose ICS following GINA guidelines, or had poorer disease control, had a higher risk of cardiovascular events following COVID-19 compared to those with moderate disease severity. Findings from a post-hoc analysis of the SUMMIT trial found that COPD patients who exacerbated had a higher risk of a subsequent cardiovascular event which remained elevated for up to a year following an exacerbation (33). This was also seen in a post-hoc analysis of the IMPACT trial (34). Given the previous literature, the findings from our study suggest that the higher risk of various post-COVID-19 cardiovascular events in people with COPD who exacerbate is likely due to COPD severity, not COVID-19. Similarly, it is well known that risk of cardiovascular disease is higher in people with asthma compared with the general population (11). Studies have found that risk factors related to age and airway obstruction were strong predictors of cardiovascular disease in people with asthma as well as COPD (35). Therefore, the association between asthma related risk factors and risk of cardiovascular events following COVID-19 in our study is likely due to the underlying respiratory disease.

This is the first study to investigate the association between pre-existing chronic respiratory disease and risk of a wide range of cardiovascular outcomes in a population of people with COVID-19 using data on the English population (20). Furthermore, due to multiple testing, we introduced a Bonferroni correction to reduce possible bias. Despite this, limitations exist. Firstly, we excluded people who had a COVID-19 vaccination prior to their first COVID-19 diagnosis which decreased the number of individuals included in the study. We excluded these individuals as it is known that the risk of post-COVID-19 outcomes and complications are modified by the COVID-19 vaccine. Previous studies have found an association between the COVID-19 vaccine and a higher risk of myocarditis and pericarditis however, we did not separate cardiovascular outcomes in our COVID-19 vaccine analysis due to low event numbers and reduced power in this analysis. Similarly, with the asthma and COPD-specific analyses, cardiovascular events were grouped together by arterial, venous, or other cardiovascular-related events due to rare events and low numbers. Results where a higher risk was seen for the composite cardiovascular outcome but not in any cardiovascular sub-group should be interpreted with caution for this reason. Lastly, residual confounding may exist as we were unable to adjust for spirometry for the COPD and asthma-specific analyses due to data availability.

## Conclusion

Using data on over three million people with COVID-19 in England, we found that people with pre-existing chronic respiratory disease had a modest higher risk of heart failure and pulmonary embolism compared with people with no pre-existing respiratory disease, but the risk might be explained at least in part by the underlying respiratory condition and severity of that condition given previous studies. COVID-19 vaccination reduced the risk of cardiovascular events following COVID-19, regardless of whether people had pre-existing chronic respiratory disease.

## Data Availability

The data used in this study are available in NHS England trusted research environment (TRE) for England, but as restrictions apply, they are not publicly available (https://digital.nhs.uk/coronavirus/coronavirus-data-services-updates/trusted-research-environment-service-for-england).

## Acknowledgments

This work is carried out with the support of the BHF Data Science Centre led by HDR UK (BHF Grant no. SP/19/3/34678). This study makes use of de-identified data held in NHS England’s TRE for England, and made available via the BHF Data Science Centre’s CVD-COVID-UK/COVID-IMPACT consortium. This work uses data provided by patients and collected by the NHS as part of their care and support. We would also like to acknowledge all data providers who make health relevant data available for research and patient and public involve

## Patient involvement

Patients were involved in the design and conduct of this research. During the grant writing stage, priority of the research question and choice of outcome measures were informed by discussions with patients through HDRUK Breathe

## Data availability and ethical approval

The data used in this study are available in NHS England’s TRE for England, but as restrictions apply, they are not publicly available (https://digital.nhs.uk/coronavirus/coronavirus-data-services-updates/trusted-research-environment-service-for-england). The CVD-COVID-UK/COVID-IMPACT programme led by the BHF Data Science Centre (https://www.hdruk.ac.uk/helping-with-health-data/bhf-data-science-centre/) received approval to access data in NHS England’s TRE for England from the Independent Group Advising on the Release of Data (IGARD) (https://digital.nhs.uk/about-nhs-digital/corporate-information-and-documents/independent-group-advising-on-the-release-of-data) via an application made in the Data Access Request Service (DARS) Online system (ref. DARS-NIC-381078-Y9C5K) (https://digital.nhs.uk/services/data-access-request-service-dars/dars-products-and-services). The CVD-COVID-UK/COVID-IMPACT Approvals & Oversight Board (https://www.hdruk.ac.uk/projects/cardiovasculard-covid-uk-project/) subsequently granted approval to this project to access the data within NHS England’s TRE for England. The de-identified data used in this study were made available to accredited researchers only. Those wishing to gain access to the data should contact in the first instance.

The Northeast - Newcastle and North Tyneside 2 research ethics committee provided ethical approval for the CVD-COVID-UK/COVID-IMPACT research programme (REC No 20/NE/0161) to access, within secure TREs, unconsented, whole-population, de-identified data from EHR data collected as part of patients’ routine healthcare.

## Author contributions

HW, AW, SW & JQ conceptulised and designed the study. HW & TB curated the data. HW & CK analysed the data. AW & JQ reviewed the methodology, data curation, data analysis and writing (original & editing). HW wrote the original manuscript. All authors reviewed and edited the manuscript.

