## Supplement p.1 for "Risk of cardiovascular events following COVID-19 in people with and without pre-existing chronic respiratory disease"

Supplementary material

### Additional methods

Baseline covariates were defined prior to the index date. These included age at index date, closest recorded body mass index (BMI) (categorised into obese and non-obese), closest recorded Index of Multiple Deprivation score (in deciles), closest recorded smoking status (never, ex-smokers, current smokers), any surgery three months prior to the index date, and history of hypertension, diabetes, cancer, liver disease, dementia, chronic kidney disease, and depression. Cardiovascular related medication, including antiplatelet, anti-lipid lowering, and anticoagulant medication, were defined as evidence of a prescription in the 3 months prior to the index date.

Due to multiple testing, Bonferroni correction was applied whereby for each set of models. For the main analysis, a p-value threshold of 0.005 was used to define statistically significant results. This was determined by dividing the p-value threshold of 0.05 by 10 for the total number of outcomes (composite cardiovascular outcome and 9 individual cardiovascular outcomes) for each cohort of people investigated. A p-value threshold of 0.0125 was used to define statistically significant results for analyses in the asthma and COPD cohort. This was determined by dividing the p-value threshold of 0.05 by 4 for the total number of outcomes (composite cardiovascular disease, arterial events, venous events, other non-arterial or non-venous events).

### Table E1: Data sources used in the English TRE

| **Data source** | **Data source name and description** |
| --- | --- |
| Primary care data | General Practice Extraction Service (GPES) data for pandemic planning and research (GDPPR) |
| Hospital episodes data | Hospital Episode Statistics (HES) and Secondary Uses Service (SUS), including admitted patient care data, outpatient data, critical care data, and Emergency Care Dataset (ECDS) |
| Mortality data | Death registry data from the Office for National Statistics (ONS) |
| Community dispensing data | NHS Business Services Authority (BSA) community dispensing data |
| COVID-19 laboratory test data | Public Health England (PHE) Second Generation Surveillance System (SGSS) test data results for COVID-19 (pillars 1 and 2) |
| COVID-19 vaccination data | National immunisation management system COVID-19 vaccination dataset |

*Legend: TRE (trusted research environment)*

### Table E2: Number of cardiovascular outcome events during follow-up.

| **Cardiovascular outcome** | **n (%)** |
| --- | --- |
| Composite cardiovascular events | 46,255 (1.26) |
| Venous thromboembolic events | 2,190 (0.06) |
| Coagulopathy events | 800 (0.02) |
| Heart failure | 14,290 (0.39) |
| Angina | 5,445 (0.15) |
| Myocarditis and pericarditis | 210 (0.01) |
| Stroke | 12,380 (0.34) |
| Myocardial infarction | 5,810 (0.16) |
| Pulmonary and arterial embolism | 4,725 (0.13) |
| Arrythmias | 420 (0.01) |

*Legend: Venous thromboembolic events include deep vein and portal vein thrombosis. Coagulopathy events include thrombocytopenia, thrombophilia and mesenteric thrombus. Heart failure includes cardiomyopathy. All numbers are rounded to the nearest 5.*

### Table E3: Baseline characteristics of people included in the study and by pre-existing respiratory disease prior to COVID-19 diagnosis

| **Baseline covariate** | **Total cohort N=3,670,460** | **No pre-existing respiratory disease N=3,000,405** | **Pre-existing respiratory disease N=670,055** |
| --- | --- | --- | --- |
| **Age (mean, SD)** | 43.1 (18.4) | 42.7 (18.1) | 44.6 (19.8) |
| **Female sex** | 1,982,115 (54.0) | 1,618,085 (53.9) | 364,030 (54.3) |
| **Smoking status**  Non-smoker  Ex-smoker  Current smoker  Missing | 2,150,610 (58.6) 833,550 (22.7) 534,220 (14.6) 152,075 (4.1) | 1,783,250 (59.4) 644,235 (21.5) 429,270 (14.3) 143,650 (4.8) | 367,355 (54.8) 189,315 (28.3) 104,950 (15.7) 8,430 (1.3) |
| **IMD**  1 (most deprived)  2  3  4  5   6  7  8  9  10 (least deprived)  Missing | 468,930 (14.8) 455,030 (12.4) 433,325 (11.8) 391,800 (10.7) 363,685 (9.9) 341,140 (9.3) 325,870 (8.9) 317,555 (8.7) 296,420 (8.1) 260,590 (7.1) 16,115 (0.4) | 376,165 (12.5) 371,090 (12.4) 355,485 (11.9) 321,870 (10.7)  297,990 (9.9) 279,875 (9.3) 266,585 (8.9) 259,965 (8.7) 242,775 (8.1) 213,640 (7.1) 14,970 (0.5) | 92,770 (13.9) 83,940 (12.5)  77,840 (11.6) 69,930 (10.4) 65,695 (9.8) 61,265 (9.1) 59,285 (8.9) 57,590 (8.6) 53,645 (8.0) 46,950 (7.0) 1,145 (0.2) |
| **Region**  Southeast  Southwest  London  East Midlands   West Midlands  East of England  Yorkshire and The Humber  Northeast  Northwest  Missing | 455,290 (12.4) 181,690 (5.0)  638,670 (17.4) 269,940 (7.4) 477,860 (13.0) 305,840 (8.3) 394,475 (10.8)  162,560 (4.4) 580,275 (15.8) 203,855 (5.6) | 373,910 (12.5) 146,180 (4.9) 543,805 (18.1) 219,730 (7.3) 386,100 (12.9) 250,270 (8.3) 318,510 (10.6) 128,925 (4.3) 465,435 (15.5) 167,545 (5.6) | 81,380 (12.2) 35,515 (5.3) 94,866 (14.2) 50,210 (7.5) 91,760 (13.7) 55,570 (8.3) 75,964 (11.3) 33,640 (5.0) 114,840 (17.1) 36,310 (5.4) |
| **BMI**  Obese  Not obese  Missing | 882,560 (24.0) 2,300,165 (62.7) 487,735 (13.3) | 682,285 (22.7) 1,882,740 (62.8) 435,375 (14.5) | 200,275 (29.9) 417,425 (62.3) 52,360 (7.8) |
| **Previous surgery** | 390,855 (10.7) | 298,345 (9.9) | 92,515 (13.8) |
| **Hypertension** | 754,915 (20.6) | 582,365 (19.4) | 172,550 (25.8) |
| **Diabetes** | 320,765 (8.7) | 241,460 (8.1) | 79,305 (11.8) |
| **Cancer** | 557,075 (15.2) | 447,130 (14.9) | 109,945 (16.4) |
| **Liver disease** | 12,185 (0.3) | 8,855 (0.3) | 3,330 (0.50) |
| **Chronic kidney disease** | 252,660 (6.9) | 185,680 (6.2) | 66,980 (10.0) |
| **Dementia** | 101,130 (2.8) | 78,870 (2.6) | 22,260 (3.3) |
| **Previous severe cardiovascular** **events** | 344,445 (9.4) | 250,725 (8.4) | 93,725 (14.0) |
| **Cardiovascular-related medication** | 797,235 (21.7) | 614,915 (20.5) | 182,322 (27.2) |

*Legend: Numbers are n(%) for binary variables and mean (SD) for continuous variables. SD (standard deviation), BMI (Body Mass Index), IMD (Index of Multiple Deprivation). All numbers are rounded to the nearest 5.*

### Table E4: Cox regression estimates (hazard ratios) for the association between baseline chronic respiratory disease and risk of CV outcomes post-COVID-19 infection -unadjusted and fully adjusted for all tables

|  | Number of CV events (n, %) | |  | | | |
| --- | --- | --- | --- | --- | --- | --- |
| CV outcome | Pre-existing respiratory disease | No pre-existing respiratory disease | Crude HR (95% CI) | P value | Adjusted HR (95% CI) | P value |
| Composite CV | 12,425 (1.86) | 33,820 (1.13) | 1.72 (1.68-1.76) | <0.0001* | 1.06 (1.03-1.08) | <0.0001* |
| VTE | 525 (0.08) | 1,645 (0.06) | 1.47 (1.34-1.62) | <0.0001* | 1.11 (1.00-1.24) | 0.0453 |
| Thrombo | 205 (0.03) | 590 (0.02) | 1.63 (1.39-1.91) | <0.0001* | 1.04 (0.87-1.24) | 0.6665 |
| HF | 4,450 (0.66) | 9,835 (0.33) | 2.12 (2.04-2.20) | <0.0001* | 1.17 (1.13-1.22) | <0.0001* |
| Angina | 1,540 (0.23) | 3,905 (0.13) | 1.86 (1.75-1.97) | <0.0001* | 1.09 (1.02-1.15) | 0.0098 |
| Myo/pericarditis | 40 (0.01) | 165 (0.01) | 1.21 (0.86-1.69) | 0.271 | 1.11 (0.77-1.59) | 0.5797 |
| Stroke | 2,845 (0.42) | 9,535 (0.32) | 1.39 (1.33-1.45) | <0.0001* | 0.88 (0.84-0.92) | <0.0001* |
| MI | 1,550 (0.23) | 4,260 (0.14) | 1.69 (1.59-1.79) | <0.0001* | 1.03 (0.97-1.09) | 0.3804 |
| PE | 1,170 (0.17) | 3,555 (0.12) | 1.52 (1.42-1.62) | <0.0001* | 1.20 (1.12-1.29) | <0.0001* |
| Arrythmia | 105 (0.02) | 310 (0.01) | 1.61 (1.28-2.03) | <0.0001* | 1.06 (0.83-1.35) | 0.6612 |

*Legend: Venous thromboembolic events include deep vein and portal vein thrombosis. Coagulopathy events include thrombocytopenia, thrombophilia and mesenteric thrombus. CV (cardiovascular), PE (pulmonary embolism).*

### Table E5: Cox regression estimates (hazard ratios) for the association between baseline chronic respiratory disease and risk of CV outcomes post-COVID-19 infection excluding people with a history of severe CV

|  | Number of CV events (n, %) | |  |  |  |  |
| --- | --- | --- | --- | --- | --- | --- |
| CV outcome | Pre-existing respiratory disease | No pre-existing respiratory disease | Crude HR (95% CI) | P value | Adjusted HR (95% CI) | P value |
| Composite CV | 6,345 (1.03) | 18,830 (0.66) | 1.62 (1.57-1.67) | <0.0001* | 1.11 (1.07-1.14) | <0.0001* |
| VTE | 365 (0.06) | 1,235 (0.04) | 1.42 (1.26-1.59) | <0.0001* | 1.18 (1.04-1.33) | 0.0101 |
| Thrombo | 135 (0.02) | 430 (0.02) | 1.51 (1.25-1.83) | <0.0001* | 1.03 (0.84-1.28) | 0.7556 |
| HF | 2,305 (0.38) | 5,485 (0.19) | 2.02 (1.92-2.13) | <0.0001* | 1.15 (1.09-1.21) | <0.0001* |
| Angina | 740 (0.12) | 2,080 (0.07) | 1.72 (1.58-1.88) | <0.0001* | 1.07 (0.97-1.17) | 0.1613 |
| Myo/pericarditis | 35 (0.01) | 150 (0.01) | 1.13 (0.79-1.63) | 0.4984 | 1.01 (0.68-1.50) | 0.9714 |
| Stroke | 1,130 (0.18) | 4,180 (0.15) | 1.29 (1.21-1.38) | <0.0001* | 0.97 (0.90-1.04) | 0.3635 |
| MI | 675 (0.11) | 2,105 (0.07) | 1.54 (1.41-1.68) | <0.0001* | 1.10 (1.00-1.21) | 0.0404 |
| PE | 895 (0.15) | 2,960 (0.10) | 1.44 (1.33-1.55) | <0.0001* | 1.20 (1.11-1.30) | <0.0001* |
| Arrythmia | 65 (0.01) | 200 (0.01) | 1.60 (1.20-2.12) | 0.0014* | 1.11 (0.82-1.52) | 0.4942 |

*Legend: Venous thromboembolic events include deep vein and portal vein thrombosis. Coagulopathy events include thrombocytopenia, thrombophilia and mesenteric thrombus. CV (cardiovascular), PE (pulmonary embolism).*

### Table E6: Cox regression estimates (hazard ratios) for the association between baseline chronic respiratory disease and risk of CV outcomes post-COVID-19 infection in those who were hospitalised for COVID-19

|  | **Complete population** | | **No history of severe CV disease** | |
| --- | --- | --- | --- | --- |
| CV outcome | Adjusted HR (95% CI) | P value | Adjusted HR (95% CI) | P value |
| Composite CV | 0.94 (0.90-0.98) | 0.0021 | 0.93 (0.88-0.98) | 0.0061 |
| VTE | 0.85 (0.70-1.04) | 0.1084 | 0.94 (0.74-1.19) | 0.6002 |
| Thrombo | 0.72 (0.52-1.00) | 0.0510 | 0.74 (0.50-1.11) | 0.1418 |
| HF | 1.09 (1.02-1.16) | 0.0064 | 1.01 (0.93-1.09) | 0.8739 |
| Angina | 0.97 (0.86-1.10) | 0.6386 | 0.91 (0.76-1.10) | 0.3224 |
| Myo/pericarditis | 1.13 (0.52-2.49) | 0.7566 | 1.10 (0.46-2.64) | 0.8354 |
| Stroke | 0.74 (0.68-0.80) | <0.0001 | 0.79 (0.69-0.90) | 0.0005 |
| MI | 0.98 (0.88-1.10) | 0.7289 | 0.98 (0.82-1.16) | 0.7813 |
| PE | 0.98 (0.88-1.10) | 0.7947 | 1.01 (0.89-1.15) | 0.8538 |
| Arrythmia | 0.88 (0.56-1.40) | 0.6010 | 0.62 (0.32-1.20) | 0.1539 |

*Legend: Venous thromboembolic events include deep vein and portal vein thrombosis. Coagulopathy events include thrombocytopenia, thrombophilia and mesenteric thrombus. CV (cardiovascular), PE (pulmonary embolism).*

### Table E7: Cox regression estimates (hazard ratios) for the association between baseline chronic respiratory disease and risk of CV outcomes post-COVID-19 infection in those who were not hospitalised for COVID-19

|  | **Total population** | | **No history of severe CV disease** | |
| --- | --- | --- | --- | --- |
| CV outcome | Adjusted HR (95% CI) | P value | Adjusted HR (95% CI) | P value |
| Composite CV | 1.09 (1.06-1.12) | <0.0001 | 1.15 (1.11-1.19) | <0.0001 |
| VTE | 1.18 (1.05-1.34) | 0.0076 | 1.21 (1.05-1.40) | 0.0092 |
| Thrombo | 1.18 (0.96-1.44) | 0.1185 | 1.14 (0.89-1.45) | 0.3079 |
| HF | 1.18 (1.13-1.24) | <0.0001 | 1.19 (1.11-1.27) | <0.0001 |
| Angina | 1.12 (1.05-1.21) | 0.0014 | 1.12 (1.01-1.24) | 0.0366 |
| Myo/pericarditis | 1.06 (0.71-1.60) | 0.7700 | 0.95 (0.61-1.49) | 0.8203 |
| Stroke | 0.93 (0.88-0.98) | 0.0118 | 1.02 (0.93-1.11) | 0.6980 |
| MI | 1.03 (0.96-1.11) | 0.3885 | 1.13 (1.00-1.26) | 0.0418 |
| PE | 1.22 (1.12-1.34) | <0.0001 | 1.19 (1.07-1.31) | 0.0009 |
| Arrythmia | 1.12 (0.84-1.47) | 0.4412 | 1.29 (0.92-1.82) | 0.1383 |

*Legend: Venous thromboembolic events include deep vein and portal vein thrombosis. Coagulopathy events include thrombocytopenia, thrombophilia and mesenteric thrombus. CV (cardiovascular), PE (pulmonary embolism).*

### Table E8: Adjusted Cox regression estimates (hazard ratios) for the association between increasing vaccine number and risk of CV outcomes post-COVID-19 infection in all patients, patients with pre-existing chronic respiratory disease, and patients with no pre-existing respiratory disease

|  | All patients | | Patients with pre-existing respiratory disease | | Patients with no pre-existing respiratory disease | |
| --- | --- | --- | --- | --- | --- | --- |
| Composite CV | Adjusted HR (95% CI) | P value | Adjusted HR (95% CI) | P value | Adjusted HR (95% CI) | P value |
| Unvaccinated period | Reference |  | Reference |  | Reference |  |
| Dose 1 | 0.57 (0.56-0.58) | <0.001 | 0.59 (0.56-0.61) | <0.001 | 0.56 (0.55-0.58) | <0.001 |
| Dose 2 | 0.56 (0.55-0.57) | <0.001 | 0.58 (0.56-0.60) | <0.001 | 0.56 (0.55-0.57) | <0.001 |
| Booster | 0.31 (0.29-0.32) | <0.001 | 0.31 (0.28-0.34) | <0.001 | 0.30 (0.29-0.32) | <0.001 |

*Legend: Reference period is the unvaccinated period from covid-19 infection to first covid-19 vaccination or censoring. The dose 1 period is from 1^st^ covid-19 vaccination to 2^nd^ covid-19 vaccination or censoring. The dose 2 period is from the 2^nd^ covid-19 vaccination to booster or censoring. The booster period is from the booster vaccine to end of follow-up or censoring. CV (cardiovascular).*

### Table E9: Risk of CVD (and each CVD component) in COVID-19 patients with pre-existing asthma compared with COVID-19 patients without asthma

|  | **Complete population** | | | | **No history of severe CV disease** | | | |
| --- | --- | --- | --- | --- | --- | --- | --- | --- |
| CV outcome | Unadjusted HR (95% CI) | P value | Adjusted HR (95% CI) | P value | Unadjusted HR (95% CI) | P value | Adjusted HR (95% CI) | P value |
| Composite CV | 1.22 (1.19-1.25) | <0.0001 | 1.04 (1.01-1.07) | 0.0026 | 1.20 (1.16-1.24) | <0.0001 | 1.08 (1.04-1.12) | <0.0001* |
| VTE | 1.26 (1.13-1.40) | <0.0001 | 1.14 (1.02-1.28) | 0.0207 | 1.25 (1.10-1.41) | 0.0005 | 1.19 (1.05-1.36) | 0.0084* |
| Thrombo | 1.12 (0.92-1.36) | 0.2507 | 0.93 (0.76-1.15) | 0.5141 | 1.11 (0.88-1.39) | 0.3678 | 0.96 (0.76-1.23) | 0.7714 |
| HF | 1.38 (1.32-1.44) | <0.0001 | 1.12 (1.07-1.17) | <0.0001 | 1.35 (1.27-1.43) | <0.0001 | 1.10 (1.03-1.17) | 0.0029* |
| Angina | 1.39 (1.29-1.49) | <0.0001 | 1.09 (1.02-1.17) | 0.0159 | 1.38 (1.25-1.52) | <0.0001 | 1.10 (1.00-1.22) | 0.0585 |
| Myo/pericarditis | 1.12 (0.78-1.60) | 0.5454 | 1.10 (0.75-1.63) | 0.6232 | 1.10 (0.75-1.61) | 0.6325 | 1.04 (0.69-1.57) | 0.8453 |
| Stroke | 0.99 (0.94-1.04) | 0.772 | 0.87 (0.83-0.92) | <0.0001 | 0.94 (0.88-1.02) | 0.1478 | 0.92 (0.85-1.00) | 0.0585 |
| MI | 1.22 (1.14-1.30) | <0.0001 | 1.05 (0.97-1.12) | 0.2224 | 1.14 (1.03-1.26) | 0.0114 | 1.09 (0.98-1.21) | 0.1116 |
| PE | 1.21 (1.12-1.30) | <0.0001 | 1.16 (1.07-1.26) | 0.0002 | 1.20 (1.10-1.30) | <0.0001 | 1.18 (1.08-1.29) | 0.0002* |
| Arrythmia | 1.26 (0.97-1.63) | 0.0811 | 1.09 (0.83-1.43) | 0.5404 | 1.28 (0.94-1.75) | 0.1201 | 1.11 (0.79-1.56) | 0.5363 |

### Table E10: Risk of CV (and subgroup) following COVID-19 in people with asthma by asthma-related risk factors

| **CV outcome** | | **Complete population** | | | | **No history of severe CV disease** | | | |
| --- | --- | --- | --- | --- | --- | --- | --- | --- | --- |
|  |  | Unadjusted HR (95% CI) | P value | Adjusted HR (95% CI) | P value | Unadjusted HR (95% CI) | P value | Adjusted HR (95% CI) | P value |
| **Asthma control** | **Composite CV** | 5.31 (5.07-5.56) | <0.001 | 1.11 (1.03-1.19) | 0.0042 | 6.12 (5.77-6.48) | <0.0001 | 1.16 (1.06-1.26) | 0.0009* |
|  | **Arterial events** | 4.83 (4.50-5.19) | <0.0001 | 1.18 (1.05-1.33) | 0.0059 | 4.70 (3.60-6.13) | <0.0001 | 1.53 (0.99-2.37) | 0.0539 |
|  | **Venous events** | 3.15 (2.80-3.55) | <0.0001 | 1.10 (0.91-1.33) | 0.3404 | 3.33 (2.88-3.84) | <0.0001 | 1.17 (0.94-1.47) | 0.1665 |
|  | **Other CV events** | 6.58 (6.16-7.03) | <0.0001 | 1.12 (1.01-1.23) | 0.0328 | 6.67 (6.24-7.12) | <0.0001 | 1.12 (1.01-1.24) | 0.0261 |
| **Asthma exacerbations** | **Composite CV** | 4.76 (4.54-5.00) | <0.0001 | 1.24 (1.15-1.34) | <0.0001 | 5.16 (4.85-5.48) | <0.0001 | 1.26 (1.15-1.38) | <0.0001* |
|  | **Arterial events** | 4.39 (5.07-4.74) | <0.0001 | 1.18 (1.05-1.33) | 0.0067 | 4.07 (3.06-5.42) | <0.0001 | 0.96 (0.62-1.48) | 0.8496 |
|  | **Venous events** | 3.02 (2.64-3.46) | <0.0001 | 1.25 (1.00-1.56) | 0.0461 | 3.06 (2.60-3.61) | <0.0001 | 1.21 (0.93-1.58) | 0.1621 |
|  | **Other CV events** | 5.63 (5.25-6.03) | <0.0001 | 1.28 (1.16-1.42) | <0.0001 | 5.71 (5.33-6.11) | <0.0001 | 1.3 (1.17-1.44) | <0.0001* |
| **ICS dose** | **Composite CV**  Low dose   Medium dose  High dose | Reference 1.60 (1.50-1.70) 2.03 (1.83-2.25) | <0.001 <0.0001 | Reference  1.08(1.01-1.15) 1.12 (1.01-1.24) | 0.0289 0.0394 | Reference  1.67 (1.54-1.81) 2.41 (2.13-2.72) | <0.0001 <0.0001 | Reference  1.09 (1.00-1.18) 1.27 (1.12-1.44) | 0.0586 0.0002* |
|  | **Arterial events**   Low dose   Medium dose  High dose | Reference 1.47 (1.33-1.64) 1.56 (1.31-1.87) | <0.0001 <0.0001 | Reference  1.03 (0.92-1.15) 0.86 (0.71-1.04) | 0.5901 0.1082 | Reference 1.73 (1.16-2.60) 2.95 (1.70-5.12) | 0.0078 0.0001 | Reference  1.16 (0.76-1.77) 1.51 (0.83-2.76) | 0.4807 0.1762 |
|  | **Venous events**   Low dose   Medium dose  High dose | Reference  1.52 (1.28-1.80) 1.57 (1.18-2.08) | <0.0001 0.0019 | Reference  1.22 (1.02-1.45) 1.02 (0.75-1.39) | 0.03 0.9095 | Reference  1.44 (1.17-1.78) 1.68 (1.20-2.36) | 0.0005 0.0027 | Reference  1.13 (0.91-1.41) 1.03 (0.71-1.49) | 0.2536 0.8766 |
|  | **Other CV events**  Low dose   Medium dose  High dose | Reference  1.70 (1.56-1.86) 2.52 (2.20-2.87) | <0.0001 <0.0001 | Reference  1.07 (0.98-1.18) 1.32 (1.15-1.52) | 0.1329 0.0001 | Reference  1.71 (1.57-1.87) 2.53 (2.21-2.88) | <0.0001 <0.0001 | Reference  1.07 (0.98-1.18) 1.29 (1.13-1.49) | 0.1444 0.0003* |

### Table E11: Risk of CVD (and each CVD component) in COVID-19 patients with pre-existing COPD compared with COVID-19 patients without COPD

|  | **Complete population** | | | | **No history of severe CV disease** | | | |
| --- | --- | --- | --- | --- | --- | --- | --- | --- |
| CV outcome | Unadjusted HR (95% CI) | P value | Adjusted HR (95% CI) | P value | Unadjusted HR (95% CI) | P value | Adjusted HR (95% CI) | P value |
| Composite CV | 8.00 (7.69-8.32) | <0.0001 | 1.09 (1.06-1.13) | <0.0001 | 8.68 (8.25-9.13) | <0.0001 | 1.17 (1.11-1.22) | <0.0001 |
| VTE | 4.29 (3.62-5.07) | <0.0001 | 1.01 (0.84-1.22) | 0.9127 | 4.48 (3.60-5.57) | <0.0001 | 1.14 (0.90-1.45) | 0.2668 |
| Thrombo | 7.48 (6.01-9.31) | <0.0001 | 1.22 (0.95-1.56) | 0.121 | 8.28 (6.24-10.98) | <0.0001 | 1.22 (0.88-1.69) | 0.2362 |
| HF | 11.43 (10.82-12.08) | <0.0001 | 1.27 (1.20-1.33) | <0.0001 | 13.54 (12.57-14.58) | <0.0001 | 1.24 (1.15-1.33) | <0.0001 |
| Angina | 8.13 (7.47-8.85) | <0.0001 | 1.08 (0.99-1.18) | 0.0859 | 7.89 (6.91-9.01) | <0.0001 | 0.97 (0.84-1.11) | 0.6275 |
| Myo/pericarditis | 2.35 (1.15-4.80) | 0.0188 | 1.37 (0.61-3.08) | 0.4508 | 2.21 (0.91-5.41) | 0.0812 | 1.06 (0.37-3.01) | 0.9145 |
| Stroke | 6.03 (5.66-6.43) | <0.0001 | 0.86 (0.80-0.92) | <0.0001 | 6.70 (6.06-7.41) | <0.0001 | 1.05 (0.93-1.17) | 0.4378 |
| MI | 7.31 (6.71-7.97) | <0.0001 | 1.02 (0.93-1.11) | 0.7368 | 8.07 (7.06-9.22) | <0.0001 | 1.19 (1.03-1.37) | 0.0204 |
| PE | 5.17 (4.65-5.75) | <0.0001 | 1.34 (1.19-1.51) | <0.0001 | 5.25 (4.58-6.00) | <0.0001 | 1.29 (1.11-1.49) | 0.001 |
| Arrythmia | 7.78 (5.79-10.46) | <0.0001 | 1.30 (0.92-1.84) | 0.1355 | 7.60 (4.96-11.65) | <0.0001 | 1.24 (0.75-2.05) | 0.4013 |

### Table E12: Risk of CV (and subgroup) following COVID-19 in people with COPD by COPD-related risk factors

| **CV outcome** | | **Complete population** | | | | **No history of severe CV disease** | | | |
| --- | --- | --- | --- | --- | --- | --- | --- | --- | --- |
|  |  | Unadjusted HR (95% CI) | P value | Adjusted HR (95% CI) | P value | Unadjusted HR (95% CI) | P value | Adjusted HR (95% CI) | P value |
| **ICS use** | **Composite CV** | 1.13 (1.05-1.21) | 0.0005 | 0.95 (0.85-1.05) | 0.2838 | 1.23 (1.13-1.33) | <0.0001 | 1.00 (0.87-1.13) | 0.9433 |
|  | **Arterial events** | 0.96 (0.88-1.06) | 0.4616 | 0.86 (0.76-0.97) | 0.0116 | 1.07 (0.71-1.61) | 0.7609 | 0.90 (0.57-1.42) | 0.6376 |
|  | **Venous events** | 1.28 (1.08-1.52) | 0.0041 | 1.15 (0.88-1.50) | 0.3045 | 1.36 (1.12-1.66) | 0.0025 | 1.33 (0.97-1.83) | 0.0786 |
|  | **Other CV events** | 1.21 (1.12-1.32) | <0.0001 | 0.99 (0.89-1.09) | 0.7779 | 1.21 (1.11-1.32) | <0.0001 | 0.98 (0.89-1.08) | 0.668 |
| **COPD exacerbations** | **Composite CV** | 2.39 (2.25-2.54) | <0.0001 | 1.40 (1.28-1.52) | <0.0001 | 2.64 (2.45-1.84) | <0.0001 | 1.52 (1.37-1.70) | <0.0001* |
|  | **Arterial events** | 2.02 (1.85-2.21) | <0.0001 | 1.14 (1.00-1.32) | 0.0564 | 1.84 (1.38-2.46) | <0.0001 | 0.96 (0.58-1.60) | 0.8781 |
|  | **Venous events** | 1.67 (1.38-2.01) | <0.0001 | 1.67 (1.24-2.24) | 0.0008 | 1.65 (1.32-2.06) | <0.0001 | 1.63 (1.14-2.33) | 0.0078* |
|  | **Other CV events** | 2.75 (2.54-2.98) | <0.0001 | 1.53 (1.36-1.72) | <0.0001 | 2.82 (2.61-3.05) | <0.0001 | 1.55 (1.38-1.74) | <0.0001* |

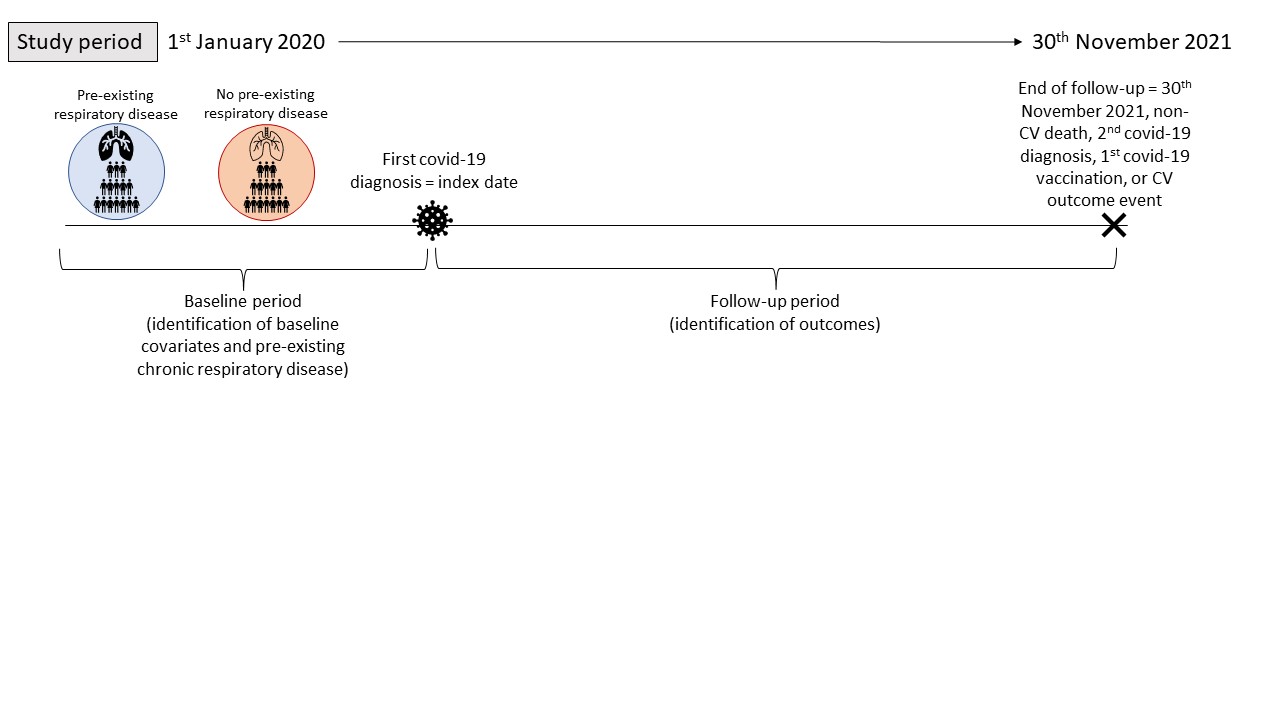

### Figure E1: Study design.

*Legend: Pre-existing respiratory disease is defined as a diagnosis of chronic obstructive pulmonary disease, asthma, bronchiectasis, cystic-fibrosis, or interstitial lung disease. CV (cardiovascular)*
